# Disruptions in Sleep Health and Independent Associations with Psychological Distress in Close Family Members of Cardiac Arrest Survivors: A Prospective Study

**DOI:** 10.1101/2024.06.18.24309137

**Authors:** Isabella M Tincher, Danielle A Rojas, Sabine Abukhadra, Christine E DeForge, Mina Yuan, S. Justin Thomas, Kristin Flanary, Daichi Shimbo, Nour Makarem, Bernard P. Chang, Sachin Agarwal

**Affiliations:** Department of Neurology, Columbia University Irving Medical Center; Wake Forest School of Medicine; Columbia University School of Nursing; Columbia University Vagelos College of Physicians and Surgeons; Department of Psychiatry & Behavioral Neurobiology, The University of Alabama at Birmingham; Cardiac arrest Family Member Stakeholder & Advocate; Division of Cardiology, Columbia University Irving Medical Center; Department of Epidemiology, Mailman School of Public Health, Columbia University Irving Medical Center; Department of Emergency Medicine, Columbia University Irving Medical Center

**Keywords:** Cardiac arrest, depression, caregiver, sleep, posttraumatic stress disorder, anxiety

## Abstract

**Background:** While recent guidelines have noted the deleterious effects of poor sleep on cardiovascular health, the upstream impact of cardiac arrest-induced psychological distress on sleep health metrics among families of cardiac arrest survivors remains unknown.

**Methods:** Sleep health of close family members of consecutive cardiac arrest patients admitted at an academic center (8/16/2021 - 6/28/2023) was self-reported on the Pittsburgh Sleep Quality Index (PSQI) scale. The baseline PSQI administered during hospitalization was cued to sleep in the month before cardiac arrest. It was then repeated one month after cardiac arrest, along with the Patient Health Questionnaire-8 (PHQ-8) to assess depression severity. Multivariable linear regressions estimated the associations of one-month total PHQ-8 scores with changes in global PSQI scores between baseline and one month with higher scores indicating deteriorations. A prioritization exercise of potential interventions categorized into family’s information and well-being needs to alleviate psychological distress was conducted at one month.

**Results:** In our sample of 102 close family members (mean age 52±15 years, 70% female, 21% Black, 33% Hispanic), mean global PSQI scores showed a significant decline between baseline and one month after cardiac arrest (6.2±3.8 vs. 7.4±4.1; p<0.01). This deterioration was notable for sleep quality, duration, and daytime dysfunction components. Higher PHQ-8 scores were significantly associated with higher change in PSQI scores, after adjusting for family members’ age, sex, race/ethnicity, and patient’s discharge disposition [β=0.4 (95% C.I 0.24, 0.48); p<0.01]. Most (n=72, 76%) prioritized interventions supporting information over well-being needs to reduce psychological distress after cardiac arrest.

**Conclusions:** There was a significant decline in sleep health among close family members of cardiac arrest survivors in the acute phase following the event. Psychological distress was associated with this sleep disruption. Further investigation into their temporal associations is needed to develop targeted interventions to support families during this period of uncertainty.

**WHAT IS KNOWN:**

- Sleep health has been identified as a key element in maintaining cardiovascular health.
- Close family members of critically ill patients experience suboptimal sleep health and psychological distress may contribute to it.

**WHAT THE STUDY ADDS:**

- It is breaking new ground in understanding the sleep health dynamics of close family members of cardiac arrest survivors, a critical but often overlooked group of caregivers.
- The study highlights significant associations between psychological distress and poor sleep that further deteriorates within the first month after a loved one’s cardiac arrest.
- Families of cardiac arrest survivors expressed a high priority for information-based interventions to help alleviate psychological distress during the initial month following the cardiac event emphasizing the need for targeted, accessible, resources to address their psychological and potentially sleep-related challenges.

## INTRODUCTION

The American Heart Association’s Life’s Essential 8 guidelines^1^ underscore the critical role of sleep in shaping cardiovascular health. This inclusion reflects a growing body of epidemiological evidence linking insufficient sleep to heightened risks of all-cause mortality and cardiovascular issues with an acknowledgment that sleep is multidimensional and other aspects of sleep health, particularly sleep quality, also contribute to cardiovascular risk.^2,3^ Close family members of critically ill patients are significantly susceptible to sleep disturbances.^4–7^ Previous investigations have highlighted poor sleep quality^4^ and daytime sleepiness^7^ in approximately half of the family members during the hospitalization of their loved one’s critical illness, with one study reporting persistent sleep disturbances among this group.^6^ While these studies examined the impact of psychological distress and feelings of uncertainty on the sleep of informal caregivers in general critical care settings, there has been limited research in the context of cardiac arrest. A recent qualitative survey study^8^ in a convenience sample shed light on disturbances in sleep among family members affected by cardiac arrest, but systematic, quantitative research remains limited.

The unique experience of cardiac arrest profoundly impacts families, especially those who witness or participate in resuscitative efforts.^9^ Close family members often endure psychological distress at levels equal to, if not greater than, survivors,^10–12^ owing to the suddenness of the event, fear of recurrence, and ensuing life changes. Moreover, the persistent uncertainty^9,13,14^ surrounding prognosis and recovery exacerbates long-term mental health challenges, including depression, generalized anxiety, and post-traumatic stress disorder (PTSD), persisting even a year after the event in some family members.^14^ Given the predictive value of distress and poor sleep for poor post-cardiac events prognosis,^15^ understanding their interplay is paramount. The bidirectional relationship between poor sleep and distress^16–18^ necessitates holistic interventions targeting both aspects. Understanding the interplay between sleep patterns and distress post-cardiac events^19,20^ could inform the development and implementation of responsive sleep interventions, particularly for the families of millions of cardiac patients hospitalized annually. Moreover, sleep health’s association with psychological health^20^ and social determinants of health^21,22^ highlights its broader implications for cardiovascular health disparities. Previous studies have suggested that racial and ethnic minorities and socioeconomically disadvantaged individuals may be disproportionately affected by suboptimal sleep patterns linked to adverse health outcomes.^23,24^ To address these knowledge gaps, we conducted a prospective study among a racially and ethnically diverse group of family members of cardiac arrest survivors at a tertiary care center in New York City. Our objectives were 1) to assess changes in sleep health metrics, including sleep quality, duration, and daytime alertness one month before and after cardiac arrest, 2) to quantify the independent effects of psychological distress markers (i.e., the severity of depression [primary predictor], generalized anxiety, and PTSD) on sleep health, 3) to identify intervention targets for prioritization in this population to potentially alleviate psychological distress and enhance post-cardiac event care.

## METHODS

### Study design

An observational longitudinal cohort study.

### Participants

Participants i.e., close family members with an intimate relationship with the patient or the assigned healthcare proxy, were identified using the daily screening logs of a National Institute of Health-funded research study (R01 HL153311) recruiting consecutive cardiac arrest patients admitted between 08/16/2021 and 06/28/2023 to eight intensive care units at the Columbia University Irving Medical Center. Enrolled participants were close family members of adult patients aged 18 years or older admitted with either in-hospital or out-of-hospital cardiac arrest. Exclusion criteria applied to participants who: 1) experienced bereavement during the study period, 2) were unavailable for in-person consent during standard working hours, or 3) did not speak either English or Spanish.

### Study assessments

Eligible family members reported demographics, medical history, and psycho-social risk factors through an intake questionnaire before hospital discharge. Measures of psychological distress were assessed one month after cardiac arrest via telephone or in-person. Sleep was measured at two time points: (1) baseline, during patients’ ICU admission after cardiac arrest, and (2) one month after cardiac arrest, a time point when most patients were discharged to home or facility. All patient-related details were obtained through electronic medical records. Columbia University Institutional Review Board approved the study protocol. This study adhered to the STROBE guidelines (see **Supplementary Methods**).

### Measures

#### Sleep Health

The Pittsburgh Sleep Quality Index (PSQI), a validated 19-item questionnaire,^25^ valuated seven components of sleep health over the previous month: sleep quality, sleep latency, duration, habitual sleep efficiency, sleep disturbance, use of sleep medication, and daytime dysfunction. A baseline assessment during hospitalization was administered to assess habitual sleep during the month before the occurrence of cardiac arrest and a follow-up assessment captured the sleep health during the month after cardiac arrest (see **Supplementary Methods**).

Scores ranged from 0 (no difficulty) to 3 (severe difficulty) and the component scores were summed to produce a global score (range 0-21). The PSQI has a sensitivity of 89.6% and a specificity of 86.5% for identifying poor sleep. Higher scores indicated poorer sleep with a PSQI score >5 indicating overall poor sleep health. Participants indicated a preference to complete the PSQI in person or via telephone.^26^ The primary outcome was the change in global sleep health score from baseline to one month after cardiac arrest. A greater change indicates a worsening in the severity of sleep health. A secondary outcome was a global sleep health score one month after cardiac arrest.

#### Depression

At one month after cardiac arrest, the Patient Health Questionnaire-8 (PHQ-8) was utilized to assess depressive symptom severity in the past two weeks about an identified traumatic experience i.e., cardiac arrest of a loved one. Participants responded to the 8 Likert-type items via telephone or in-person, with responses associated with a score: “not at all” (0), “several days” (1), “more than half the days” (2), and “nearly every day” (3). The sub-scores were summed to get a final score between 0-24. The PHQ-8 has good validity and reliability for detecting major depression, with a sensitivity of 88%, a specificity of 88%, and a high Cronbach’s Alpha value (0.89).^27^

#### Generalized Anxiety

The Generalized Anxiety Disorder-2 (GAD-2) is a validated, shorter version of GAD-7, using only the first two questions to screen for general anxiety symptoms. It has excellent psychometric properties of the GAD-7 with high sensitivity (86%) and specificity (83%).^28^

#### Post-traumatic Stress

The PTSD Checklist (PCL-5) is an extensively validated, 20-item scale developed by the National Center for PTSD that corresponds to *DSM-5* criteria for PTSD. The PCL-5 has adequate test-retest reliability and excellent sensitivity/specificity for PTSD clinical diagnosis prediction.^29^

### Prioritization Exercise of Supportive Interventions

In the context of prioritizing supportive interventions post-cardiac arrest, proposed measures previously delineated^30^ were formulated via a collaborative, co-designing process involving primary stakeholders (see **Supplementary Methods**). These interventions encompass eight distinct categories, broadly classified as either information-centric (education on potential recovery, knowledge about cardiac arrest, access to care teams, understanding rehabilitation options) or well-being-centric (access to caregiver resources, self-care tools, connection with other caregivers, and professional psychotherapy). At one month after cardiac arrest, participants were described the 8 interventions and asked to assess their perceived helpfulness on a 5-point Likert scale ranging from “extremely helpful” to “not at all helpful.” Then, participants were prompted to indicate their preferred mode of intervention delivery (i.e., written materials, web-based platforms, in-person sessions, telephonic consultations, mobile/computer applications).

Lastly, they were tasked with prioritizing the top three interventions according to their perceived necessity in addressing their needs during the initial month of post-cardiac arrest.

### Statistical Analyses

Sleep health metrics between the two assessments were compared using chi-square and t-tests. The association between cardiac arrest-induced depression and sleep health was examined in an unadjusted and adjusted linear regression model with total PHQ-8 score as the primary predictor and change in global PSQI score (one-month PSQI score – baseline PSQI score) as the primary dependent variable. Following the recommended guidelines, we selected covariates a priori.^31^ Based on published findings of factors that might confound the association between psychological distress and sleep health, Model 1 included demographics (age, sex, race/ethnicity)^7,32,33^ and baseline global PSQI score, and Model 2 additionally included the patient’s discharge disposition,^6,30^ reflecting post-acute care needs.

For potential significant (p<0.01) factors to be included in Model 3, we explored independent associations of family- and patient-related factors listed in **Table 1** with the primary outcome. We considered (1) <u>family member attributes</u> (e.g., marital status, relationship to the patient), (2) <u>psychological risk factors</u> (e.g., whether family members had witnessed the cardiac arrest, factors associated with positive psychological well-being including optimism, purpose in life, pre-existing diagnosis of psychiatric disorders such as depression, anxiety or post-traumatic stress, or current use of spirituality as a coping mechanism), (3) <u>patient-related characteristics</u> (e.g., age, sex, health insurance, location of arrest, length of ICU stay), and (4) <u>hospital-related outcomes</u> (e.g., received a tracheostomy, discharge functional status).

**Table 1.**
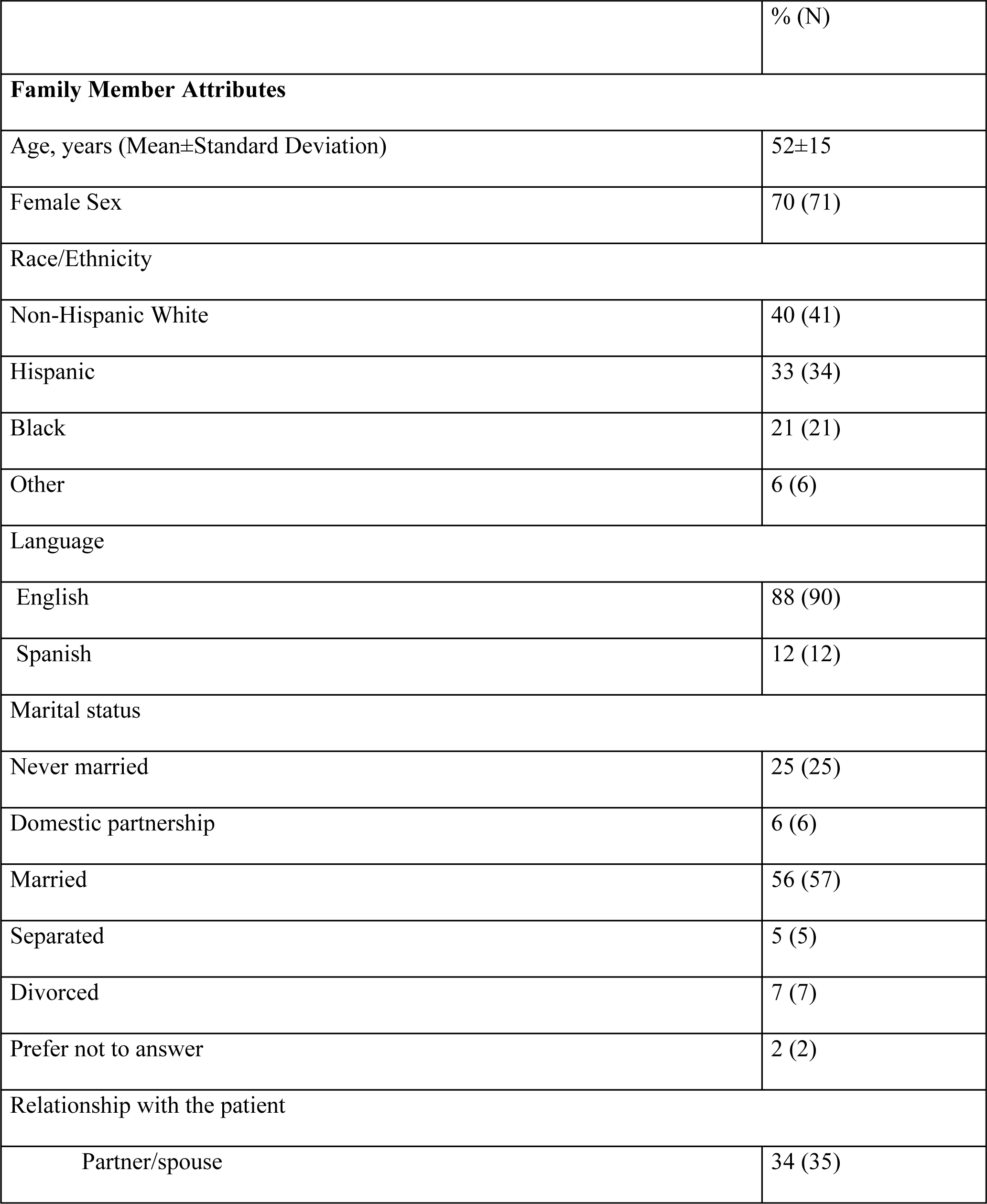
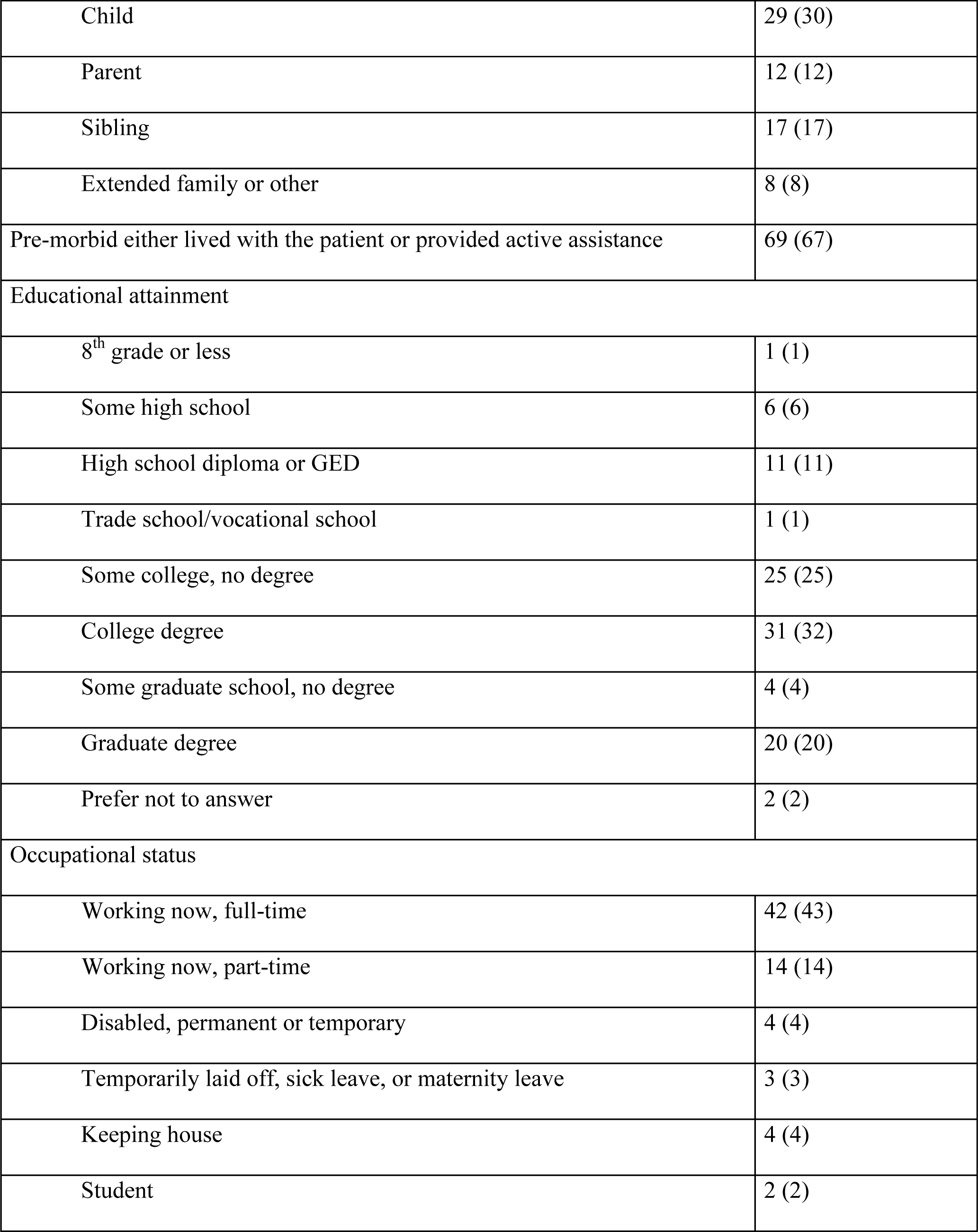
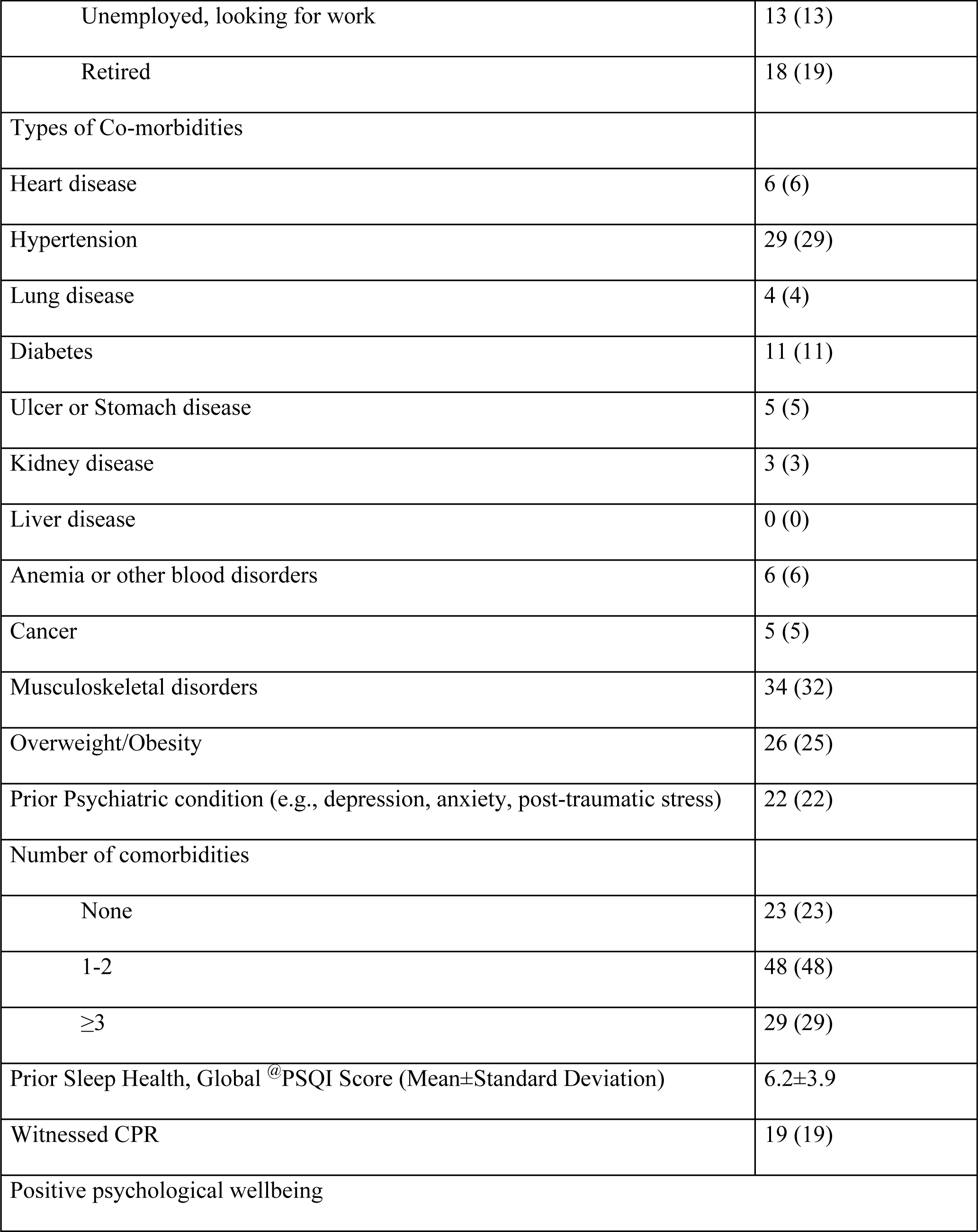
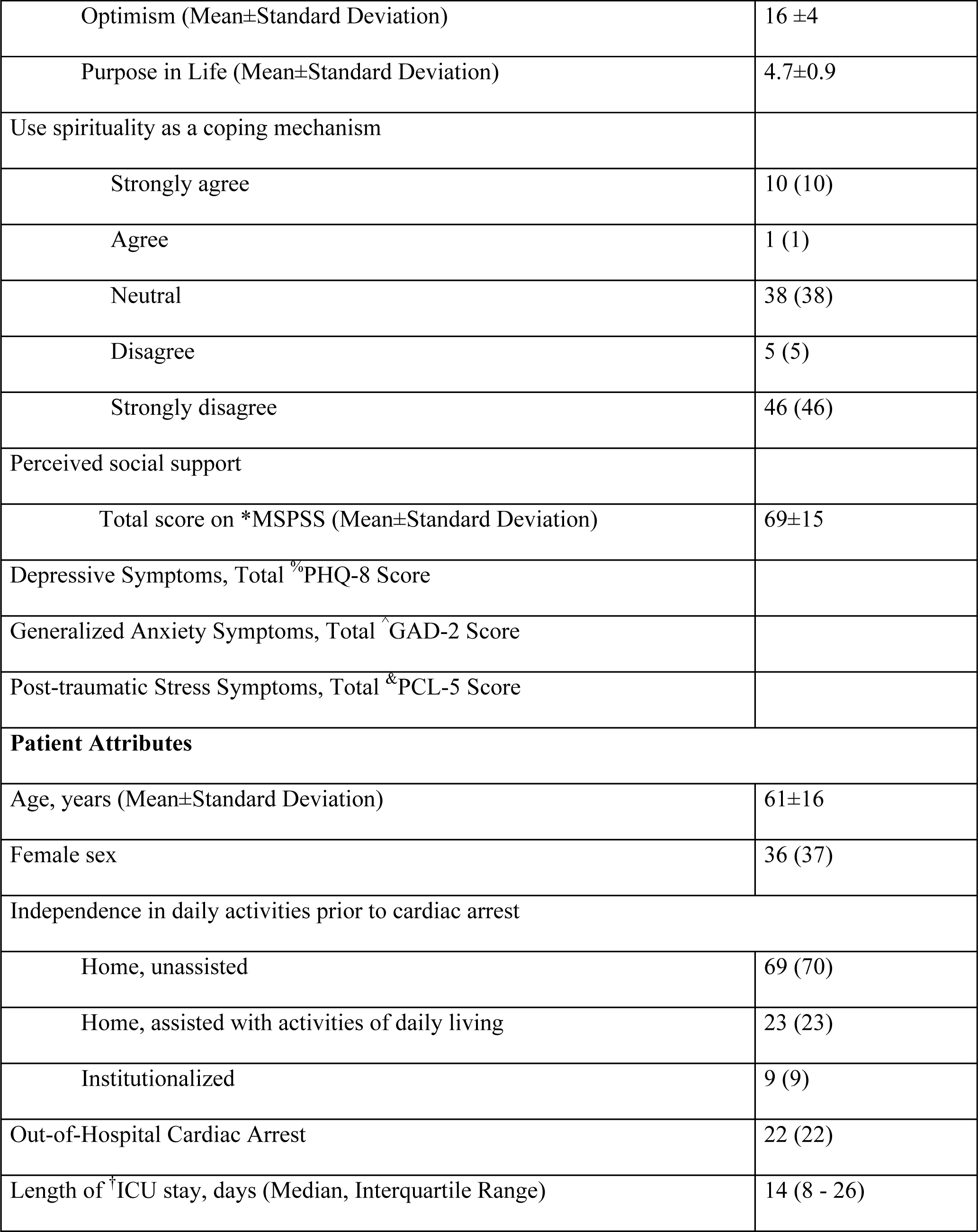
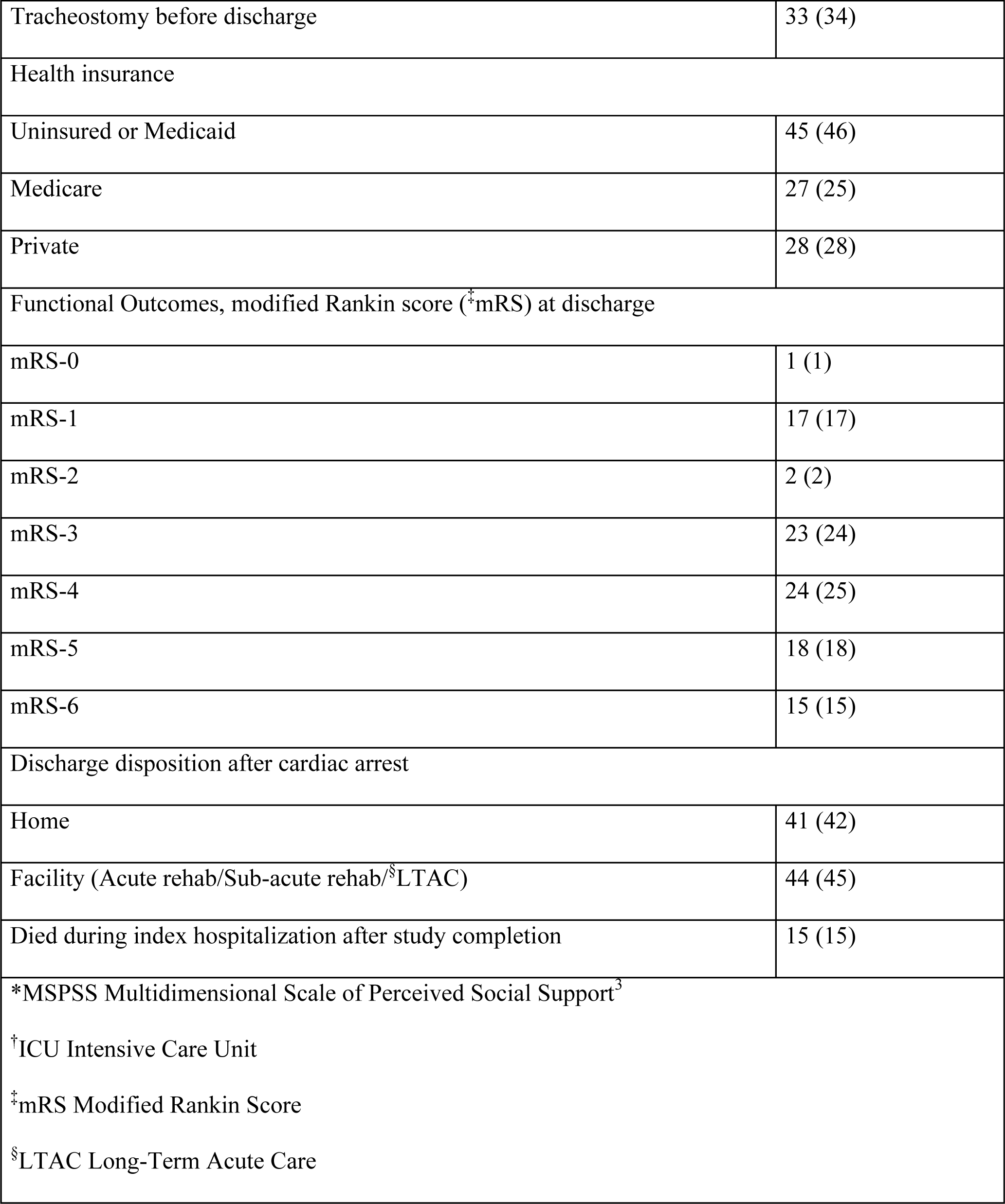
Characteristics of Family Members of Cardiac Arrest Survivors (N=102).

Similarly, two separate analyses tested the associations of GAD and PTSD total scores with differences in global PSQI scores after adjusting for the covariates included in the final model for PHQ-8.

### Post-regression analysis

After estimating the beta coefficients and their significance, to provide a comprehensive understanding of the relationships between depression and sleep health, we calculated 1) R^2^ value to indicate the overall strength of the relationship and a measure of the model’s goodness of fit. 2) Effect Size for individual predictors using eta-squared (η^2^), 3) Confidence intervals for effect sizes to convey the precision of your estimates.

### Sensitivity analysis

We repeated our primary outcome analyses with PHQ-8 total score after excluding scores obtained on the sleep-related question to prevent bias resulting in overestimation of the association.

### Power analysis

Using the R-squared test in multiple linear regression, with alpha=0.05, sample size 100, R-squared of 0.53, and 7 tested covariates, the estimated power was greater than 95%.

### Secondary Outcomes

We then conducted similar analyses of PHQ-8, GAD-2, and PCL-5 total scores with global PSQI scores at one month.

### Intervention Prioritization

Key measures and statistical analysis centered on the relative performance and ranking of the eight interventions tested. We first compared the performance based on combined ‘very helpful to extremely helpful’ responses on the Likert Scale. The top three ranks were used as the key measure for individual intervention comparisons. Further categorization into two a priori groups of interventions - ‘information needs’ vs ‘well-being needs’ was based on the top-rank assignment for individual interventions.

All statistical analysis was performed using STATA 18. A significance level of p<0.05 was considered statistically significant.

## RESULTS

### Study population

Of 438 cardiac arrest admissions, 247 met exclusions. Of 191 eligible participants approached, 137 consented and 102 with one-month outcomes data were analyzed **(CONSORT Diagram, Supplementary Figure 1)**. As shown in **Supplementary Table 1**, compared to 25 participants who either withdrew or did not complete a one-month assessment a higher proportion of the analyzed group had an education of college degree or higher (28% vs 56%) and went home (12% vs 43%). There were more deaths during hospitalization after the study duration of 1 month after cardiac arrest in the lost to follow-up group (40% vs 15% in the analyzed group).

### Close family members’ attributes

Participants’ average age was 52±15 years, with the majority identifying as female (n=71, 70%) and as Black (n=21, 21%) or Hispanic (n=34, 33%); Black race n=21, 21%). The most common relation to the patient was a spouse/partner (n=35, 34%). A few participants, 12 (12%), were Spanish-speaking only, and 19 (19%) witnessed cardiopulmonary resuscitation. Most participants reported one or more comorbid conditions (n=79, 77%), with musculoskeletal disorders (n=32, 34%), hypertension (n=29, 29%), obesity (n=25, 26%), and prior psychiatric history (n=22, 22%) being most common **(Table 1)**.

### Patient characteristics

Almost all patients lived at home before cardiac arrest (n=93, 92%). After a median (interquartile range) ICU length of stay of 14 (8-26) days, 34 (33%) required tracheostomy, 58 (57%) had poor functional status (modified Rankin score >3) at hospital discharge, and less than half (n=42, 41%) went home **(Table 1).**

### Sleep Health and its components

The mean global PSQI score increased significantly from baseline to one month after cardiac arrest (6.2±3.8 vs 7.4±4.1; p<0.01) **(Table 2).** The mean difference in PSQI scores was 1.3±3.8. Any worsening in global PSQI scores was seen in 43% (n=43) while 23% had their scores unchanged between the two assessments.

**Table 2.** Bivariate associations of various family member and patient characteristics with Differences in Sleep Health between baseline and one month after cardiac arrest (N=100).

|  | **Coefficients** | **P-value** |
| --- | --- | --- |
| **Family Member Attributes** |  |  |
| Age, years | -0.04 | 0.09 |
| Female Sex | -1.1 | 0.2 |
| Race/Ethnicity |  |  |
| Non-Hispanic White | Reference |  |
| Hispanic | 2.8 | **<0.01** |
| Black | 0.9 | 0.3 |
| Other | 1.0 | 0.5 |
| Primary Language (Spanish vs English) | 3.5 | **<0.01** |
| Marital status (Married/Domestic Partnership vs others) | -0.06 | 0.9 |
| Relationship with the patient |  |  |
| Partner/spouse | Reference |  |
| Child | 0.9 | 0.3 |
| Parent | -0.6 | 0.6 |
| Sibling | 0.5 | 0.6 |
| Extended family or other | -1.0 | 0.4 |
| Pre-morbid either lived with the patient or provided active assistance | 0.2 | 0.8 |
| Education, College degree or higher vs others | -0.6 | 0.4 |
| Occupation, Working full or part-time vs others | 0.6 | 0.4 |
| Number of comorbidities |  |  |
| None | Reference |  |
| 1-2 | 0.7 | 0.5 |
| $\geq 3$ | 0.3 | 0.8 |
| Prior Psychiatric History (Yes/No) | 0.06 | 0.9 |
| Prior Sleep Health, Global <sup>@</sup>PSQI Score | -0.4 | **<0.01** |
| Witnessed CPR of their loved one (Yes/No) | 1.3 | 0.2 |
| Positive psychological wellbeing |  |  |
| - Optimism | 0.09 | 0.2 |
| - Purpose in Life | -0.5 | 0.2 |
| Use spirituality as a coping mechanism, Disagree or Strongly Disagree vs others | 0.3 | 0.4 |
| Perceived social support, <sup>\$</sup>MSPSS Total score | -0.04 | 0.1 |
| Depressive Symptoms, Total <sup>%</sup>PHQ-8 Score | 0.4 | **<0.01** |
| Generalized Anxiety Symptoms, Total <sup>^</sup>GAD-2 Score | 0.8 | **<0.01** |
| Post-traumatic Stress Symptoms, Total <sup>&</sup>PCL-5 Score | 0.09 | **<0.01** |
| **Patient Attributes** |  |  |
| Age | 0.007 | 0.8 |
| Female sex | -0.03 | 1.0 |
| Poor Health insurance (Uninsured/Medicaid vs others) | -0.9 | 0.2 |
| Out-of-Hospital Cardiac Arrest | 0.3 | 0.7 |
| Length of <sup>†</sup>ICU stay | 0.004 | 0.8 |
| Discharged with a Tracheostomy, (Yes vs No) | -0.3 | 0.7 |
| Poor Functional Outcomes (modified Rankin score >3) at discharge | -0.2 | 0.7 |
| Discharge disposition after cardiac arrest |  |  |
| Home | reference |  |
| Facility (Acute rehab/Sub-acute rehab/<sup>§</sup>LTAC) | -3.1 | **<0.01** |
| Died during index hospitalization after study completion | -1.3 | 0.2 |
| *All associations were adjusted for baseline global PSQI score<br>@PSQI Pittsburgh Sleep Quality Index<br>\$MSPSS Multidimensional Scale of Perceived Social Support<br>%PHQ-8<br>^GAD-2<br>&PCL-5<br><sup>†</sup>ICU Intensive Care Unit<br><sup>§</sup>LTAC Long-Term Acute Care | | |

Of all PSQI components, <u>sleep quality</u>, <u>sleep duration</u>, and <u>daytime dysfunction</u> showed significant worsening from baseline to one month after cardiac arrest **(Figure 1)**. There was a significant increase in the proportion of participants reporting poor sleep quality from baseline to one month after cardiac arrest (24% vs 47%; p<0.01). Among participants with good sleep quality at baseline (76%, n=75/99), 31% (n=23/75) shifted to poor sleep quality at one month. Among participants with guideline-recommended sleep duration of ≥7 hours at baseline,^34^ 40% (n=19/48) had worsening in their sleep duration one month after cardiac arrest (10% reported sleep duration of <5 hours, 15% with ≥5 and <6 hours, 15% with ≥6 and <7 hours).

**Figure 1.**
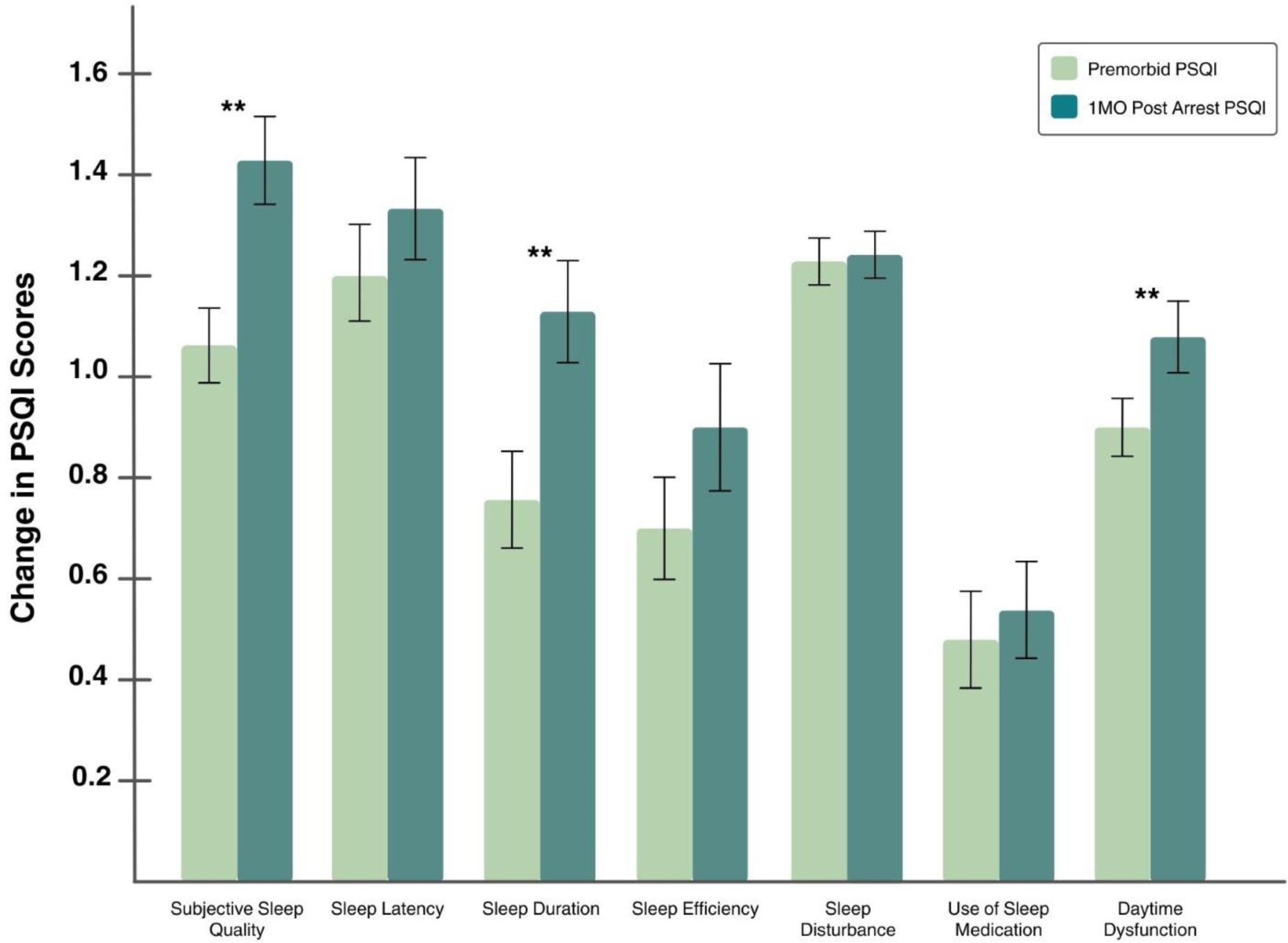
Changes in PSQI Scores of Close Family Members of Cardiac Arrest Survivors from Pre-Morbid Evaluation to 1 Month Post-Arrest by Subcategory.

### Means of depressive, generalized anxiety, and PTSD symptom scores one month after cardiac arrest

The average PHQ-8, GAD-2, and PCL-5 scores at a median duration of 28.5 (interquartile range 10-63) days after cardiac arrest were 7.0±5.5, 2.8±2.1, and 14.8±12.4, respectively.

### Covariate selection for the multivariable-adjusted model

Apart from PHQ-8, GAD-2, and PCL-5 total scores, Hispanic ethnicity, and patients’ discharge disposition to home showed significant (p<0.01) bivariate associations with differences in global PSQI scores. All associations were adjusted for baseline global PSQI score. Since there were only 12% (n=12) participants in the Spanish-speaking group, though statistically significant, it was not included in the multivariate model **(Table 2)**.

### Association of Psychological Distress with Sleep Health

Higher PHQ-8 scores were significantly associated with greater change in total global PSQI scores **(Table 3),** both in the bivariate analysis after adjusting for baseline PSQI score (β=0.4, 95% CI 0.2–0.5), p<0.01) and in a multivariate model after adjusting for age, female sex, race/ethnicity, and discharge disposition (β=0.3, 95% CI 0.2–0.4), p<0.01). The 95% confidence interval for *R*^2^ (0.53) ranged from 0.3 to 0.6, indicating a relatively precise estimate of the proportion of variance explained by the full model The effect size (η^2^) for PHQ-8 was 0.2 (95% CI 0.1–0.4), indicating a moderate effect on the sleep health **(Supplementary Table 2).** Higher GAD-2 and PCL-5 total scores were significantly associated with greater differences in global PSQI scores (β=0.6, 95% CI 0.3–0.9, p<0.01 and β=0.06, 95% CI 0.01–0.1, p<0.01, respectively) in a multivariate model **(Supplementary Tables 3 and 4)**.

**Table 3.** Associations Between Depressive Symptoms in Close Family Members and Changes in Sleep Health Before and 1 Month After Cardiac Arrest.

| Covariates | Multivariate | Coefficient | P value | Multivariate | Coefficient | P value |
| --- | --- | --- | --- | --- | --- | --- |

|  | **Model 1** | **(95% CI)** |  | **Model 2** | **(95% CI)** |  |
| --- | --- | --- | --- | --- | --- | --- |
| **PHQ total score** | 0.4 | (0.2, 0.5) | **<0.01** | 0.3 | (0.2, 0.4) | **<0.01** |
| **Age** | -0.003 | (-0.04, 0.04) | 0.8 | -0.001 | (-0.04, 0.04) | 1.0 |
| **Female sex** | -0.01 | (-1.2, 1.2) | 0.9 | 0.07 | (-1.2, 1.3) | 1.0 |
| **Non-Hispanic White** | reference |  |  | reference |  |  |
| **Hispanic** | 2.0 | (0.6, 3.3) | **<0.01** | 1.7 | (0.4, 3.1) | **0.01** |
| **Black** | 1.1 | (-0.4, 2.6) | 0.1 | 1.1 | (-0.4, 2.5) | 0.1 |
| **Other** | 1.6 | (-0.7, 4.0) | 0.1 | 1.3 | (-1.0, 3.7) | 0.2 |
| **Pre-PSQI total score** | -0.6 | (-0.7, -0.4) | **<0.01** | -0.6 | (-0.8, -0.5) | **<0.01** |
| **Home** |  |  |  | reference |  |  |
| **Facility** |  |  |  | -1.4 | (-2.7, -0.1) | **0.03** |
| **Died** |  |  |  | -0.8 | (-2.5, 0.9) | 0.4 |

### Sensitivity Analysis

Higher PHQ-8 scores even after the exclusion of values on sleep-related questions were significantly associated with greater differences in total global PSQI scores after adjusting for age, female sex, race/ethnicity, and discharge disposition [β=0.3, 95% CI 0.2 - 0.4, p<0.01]. **(Table 3)**

### Secondary Outcome

We found similar significant associations in an adjusted model of one-month PHQ-8, GAD-2, and PCL-5 total scores with global PSQI score at one month **(Supplementary Tables 5-8).**

### Prioritization Results of Supportive Interventions

In the month following the cardiac arrest, most participants (76%; n=72) preferred one of the four information-based interventions (i.e., education on potential recovery, knowledge about cardiac arrest, access to care teams, and understanding rehabilitation options) as opposed to well-being needs (24%; n=23) including access to caregiver resources, self-care tools, connection with other caregivers, and professional psychotherapy.

The top three interventions rated ‘very helpful’ or ‘extremely helpful’ as a supportive resource were access to the care team (94% of participants; n=89/95), education on potential recovery (93%; n=88/95), and understanding rehabilitation options (87%; n=83/95) **(see Figure 2).** Of 760 responses, written information (27%; n=204) and web-based information (26%; n=194) were the most preferred modes of delivery, while a phone app resource received the least preference (11%; n=83). Specifically, written information or web-based was the most preferred mode of delivery for all intervention resources except having access to the care teams and professional psychotherapy, where participants preferred phone calls **(Figure 3)**.

**Figure 2.**
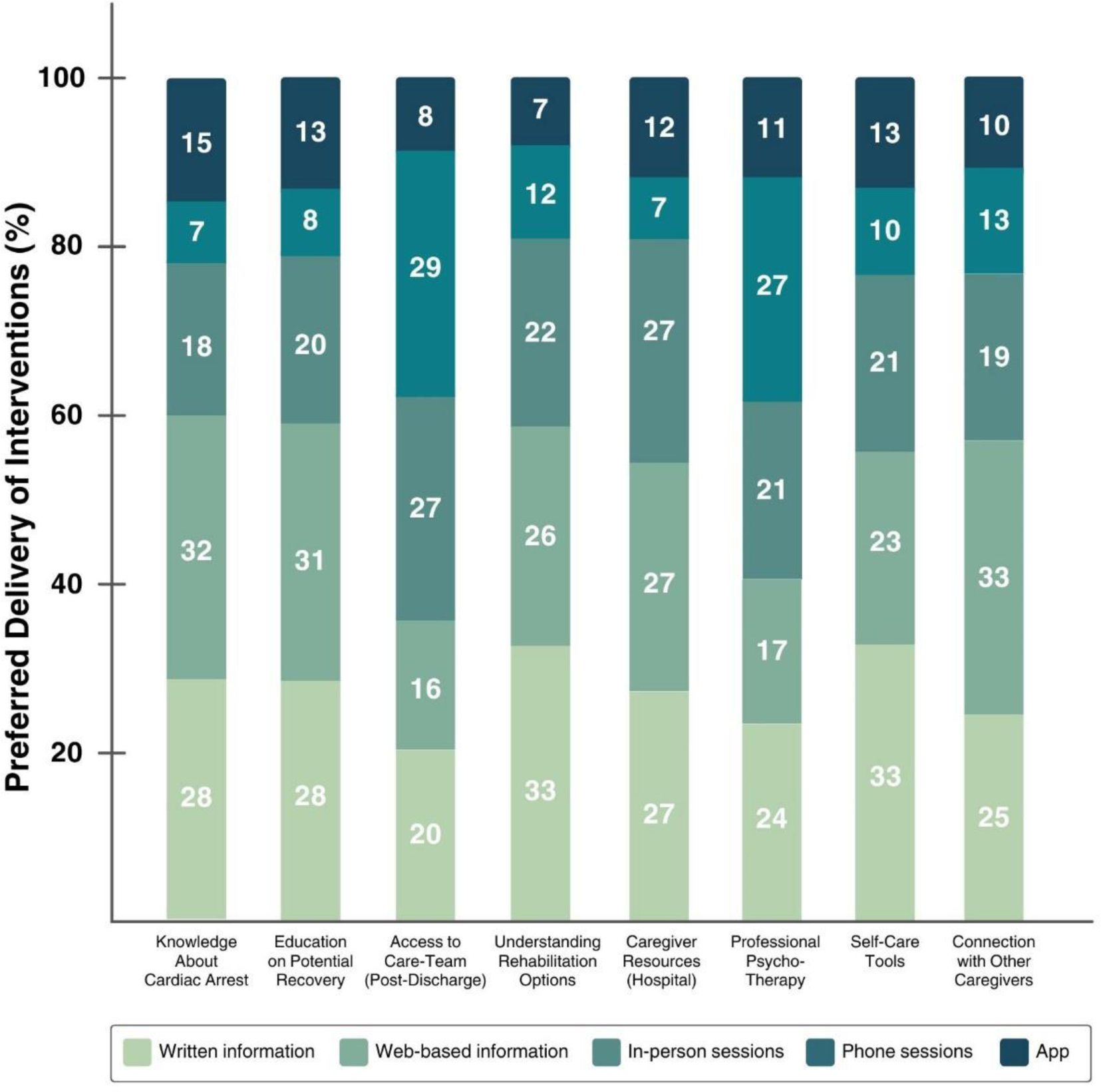
Evaluation of Preferred Method of Delivery for Interventions Aimed to Reduce Psychological Distress in Close Family Members of Cardiac Arrest Survivors.

**Figure 3.**
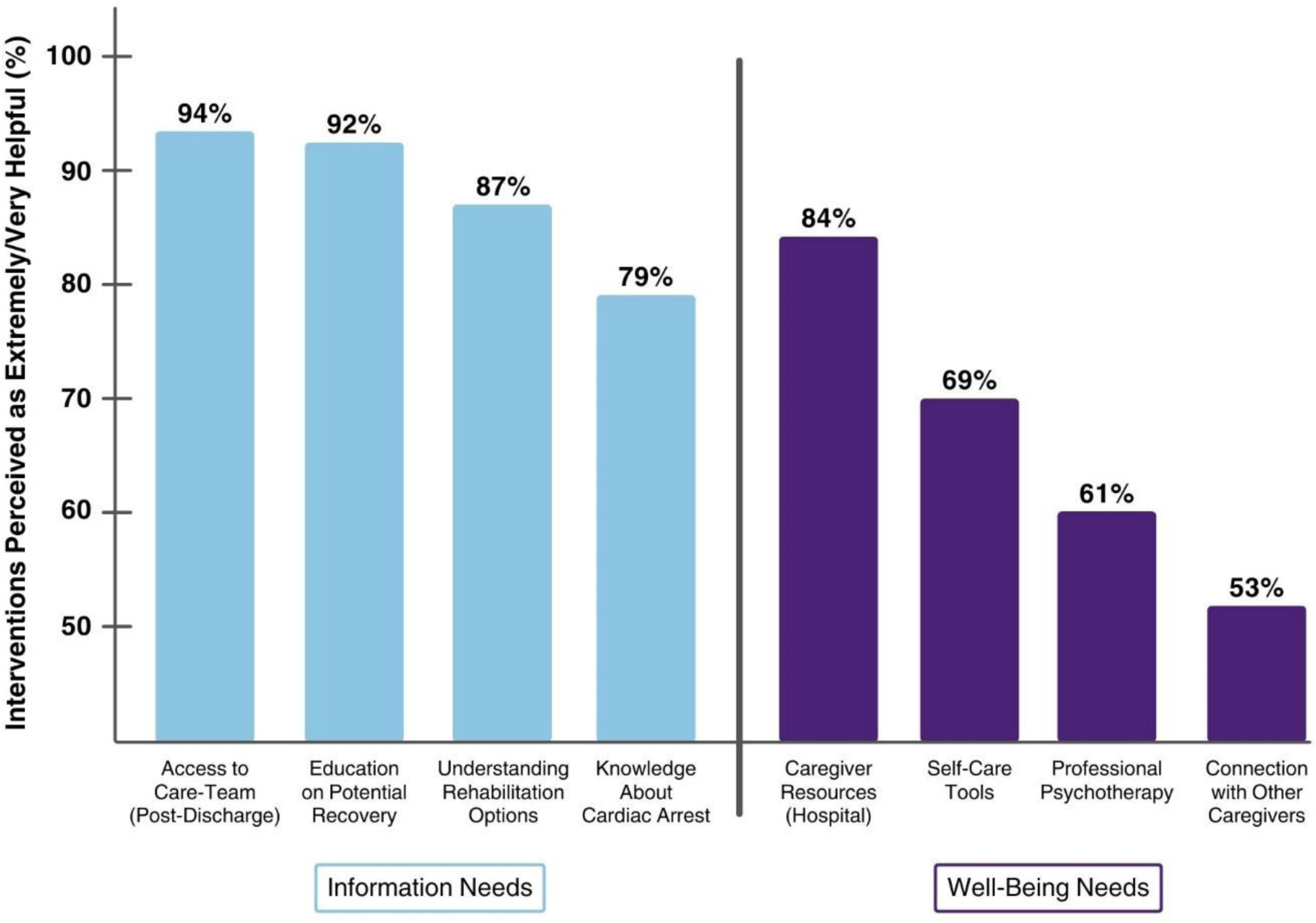
Prevalence Estimates of Depression, Generalized Anxiety, and Post-Traumatic Stress Disorder at 1 month in Close Family Members of Cardiac Arrest Survivors.

The top three interventions ranked as either first, second or third were (A) Access to the care team for information beyond hospital discharge (62%, n=59/95), (B) Education on the potential neurological, physical, and emotional recovery expected during hospitalization and up to a year after discharge (55%, n=52/95), and (C) Knowledge about cardiac arrests and their distinctions from heart attacks or strokes (41%, n=39/95) **(Figure 4).**

**Figure 4.**
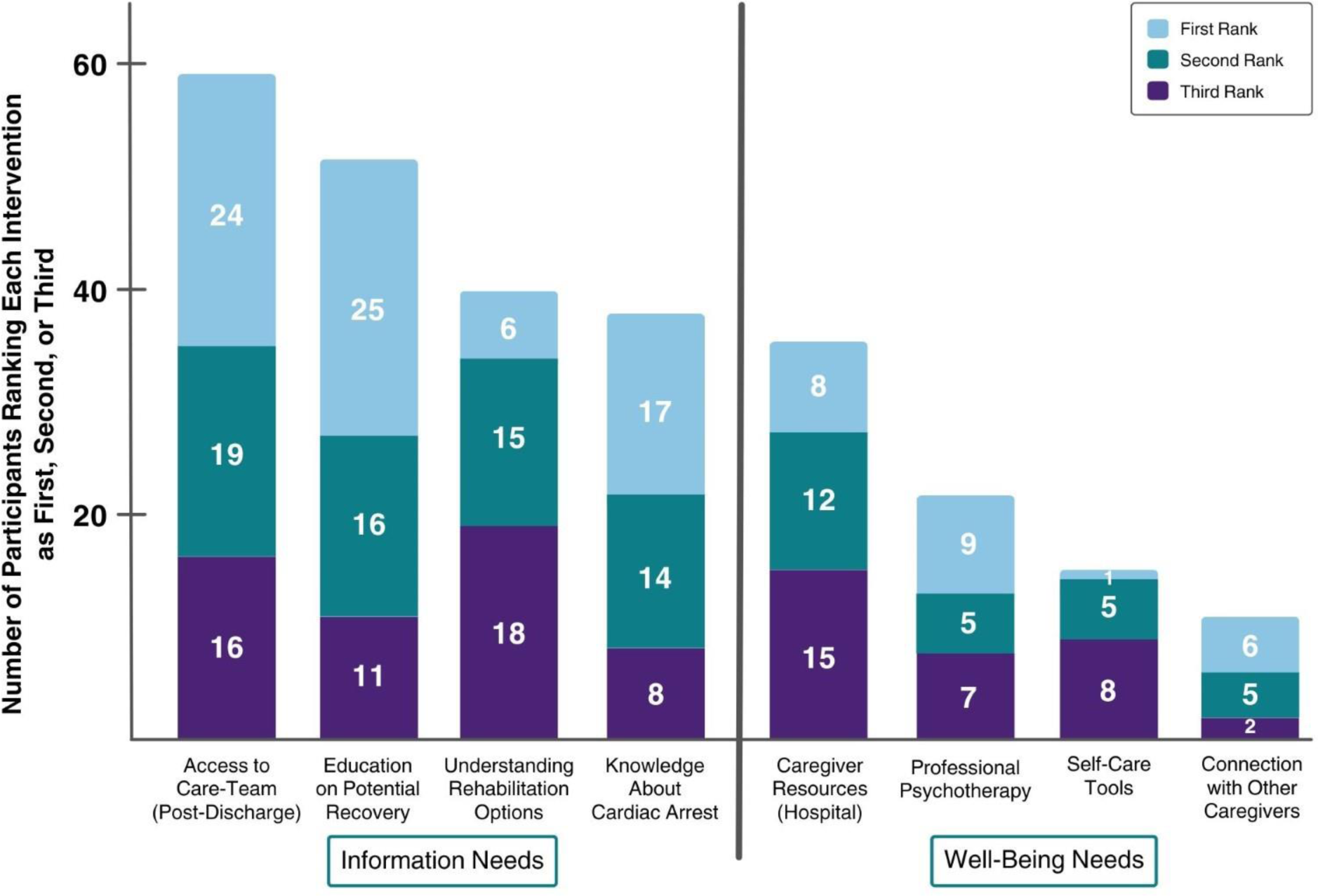
Number of Each Intervention Ranked as First, Second, or Third by Close Family Members of Cardiac Arrest Survivors.

## DISCUSSION

This study is among the earliest, longitudinal, systematic quantitative assessments of sleep health in close family members of cardiac arrest survivors. Our findings confirm that family members experience a significant decline in sleep health within the initial month after the event of their loved one. One study^6^ found that over half of families of patients on prolonged mechanical ventilation continued to experience poor sleep even two months post-discharge, highlighting the enduring nature of the experience. Understanding sleep patterns prior to distressing events can facilitate understanding of underlying factors contributing to the decline in sleep health and guide early interventions in the hospital setting.

In addition to subjective sleep quality, participants reported significant disturbances in sleep duration and daytime dysfunction sub-components of the PSQI. Among those who followed the recommended sleep duration of ≥7 hours, 40% saw worsened sleep duration one month post-event, an increase vs. 24% at baseline. This may reflect critical care experiences, where family members encounter life and death decisions both in the ICU setting and post-discharge. Our results align with previous research^6,7^ indicating that poor sleep quality at night results in excessive daytime sleepiness, impaired functionality, and cognitive clarity among family members of critically ill patients. PSQI measures daytime dysfunction by asking about enthusiasm to complete tasks and trouble staying awake. Sleep disturbances challenge caregiving and serve as a potential risk factor for post-intensive care syndrome - Family. These findings underscore the need to address sleep-related issues in family members of critically ill patients to improve outcomes and well-being.

The decline in sleep health among close family members is significantly correlated with depression symptoms within the initial month following the cardiac event. Statistically significant fully adjusted associations were also found between the deterioration of sleep and GAD and PTSD symptoms. These findings highlight the intricate relationship between psychological distress and sleep disturbances in the aftermath of sudden, traumatic, and life-altering incidents, indicating potential intervention opportunities.^8^ Our research extends the limited evidence demonstrating a high sleep disturbance prevalence and elucidates its correlation to psychological distress among family members in other critical care conditions.^6^ A previous internet-based support group qualitative study found that 95% (n=37/39) of family members experienced persistent psychological distress, with one in four reporting poor sleep following hospital discharge.^8^ Another cross-sectional study of 200 family members of critically ill patients revealed a bi-directional relationship^16^ showing significant associations between sleep disruptions during hospital stays and subsequent symptoms of depression and anxiety.^5^

One noteworthy discovery warranting discussion pertains to the susceptibility of Hispanic participants to sleep health issues compared to their non-Hispanic white counterparts, a phenomenon previously documented in community-based studies.^24,35–37^ At one month, a pronounced discrepancy in good sleep was observed between Hispanic and non-Hispanic white participants (9% vs. 56%) (refer to **Supplementary Table 9**). Additionally, Hispanic ethnicity demonstrated significant links with declining sleep health over time, in contrast to non-Hispanic whites. These associations persisted across all models for the three psychological distress indicators studied. These disparities among racial/ethnic minorities underscore the potential role of sleep, in conjunction with psychosocial factors, in contributing to observed inequities in cardiovascular health.^20^ While noteworthy sleep variations were evident among English and Spanish-speaking participants exclusively, the limited sample size (n=12) underscores the need for further exploration in larger cohorts. Intriguingly, no discernible sex-based distinctions were apparent in either sleep health or its association with psychological distress. However, considering that approximately 66% of informal caregivers in the United States are female, future investigations should prioritize examining this aspect.

Discharge to a rehabilitation facility, rather than home, after hospitalization, may protect sleep health by reducing the potential impact of anxiety, uncertainty, and unpreparedness experienced by families at the time of discharge. This aligns with a previous study^6^ showing families of patients discharged home within two weeks of ICU discharge exhibited poorer objective and subjective sleep compared to those discharged home two months post-ICU discharge. It was theorized that early home discharge may exacerbate stress by overwhelming caregivers with increased care demands, whereas discharge to a rehabilitation facility may potentially provide reassurance to families that their loved ones will continue to receive professional care. Future studies should explore whether varied post-ICU transitions influence families’ sleep to identify potential intervention targets during this vulnerable period. Our third objective investigated interventions targeting the root causes of poor sleep health among family members during the acute period. Family members prioritized information-based interventions to alleviate uncertainty and psychological distress, which in turn could potentially improve sleep health. This finding confirms the themes from a qualitative study on in-hospital cardiac arrest,^38^ a National Institute of Health funded workshop,^39^ and our previous work^30^ on families’ post-cardiac arrest experiences and needs. In our cross-sectional survey of a national sample (n=550) of close family members within six months of their loved one’s cardiac arrest, we identified factors influencing families’ preferences for information versus well-being needs including age over 40 years, witnessing the cardiac arrest, early caregiving, and direct discharge home. Interestingly, the study revealed a shift in intervention preferences over time. The need for psychological support increased significantly 2–6 months post-discharge, highlighting families’ evolving needs and the importance of ongoing support.

Another study found that 58% of family members of critically ill patients experienced moderate to severe sleep disturbances during hospitalization, with more information about their loved one’s health being the most common suggested remedy to improve their sleep.^4^ Additionally, resourceful family members reported lower levels of depression, anxiety, and sleep disturbances.^5^

Future studies might employ experimental designs to evaluate psycho-education interventions or cognitive behavioral therapy^20^ in addressing the mental and sleep health needs of family members. Targeted interventions tailored to specific needs can better support this vulnerable population throughout the recovery process.

## STRENGTHS

There are several study strengths. This investigation focused on family members of cardiac arrest survivors, a previously understudied group, providing valuable insights into their unique experiences and needs. By excluding those in bereavement, we ensured a clear focus on this population. Further, our study’s adequate power and prospective study design enhance the internal validity of our findings. The recruitment of family members from a sex and racially and ethnically diverse study population further strengthens the external validity of our results.

Our comprehensive approach analyzes both patient and family member characteristics while examining potential associations between psychological distress and sleep health. It provides a nuanced understanding of the factors influencing sleep disturbances among family members of cardiac arrest survivors.

Finally, our study introduces the novel aspect of examining sleep health decline within the first month after cardiac arrest in comparison to pre-event sleep patterns. We also investigate changes in other aspects of sleep health that are predictive of morbidity and mortality such as sleep duration, quality, and daytime dysfunction.^40^ This innovative approach sheds light on the temporal dynamics of sleep disturbances, offering valuable insights into the trajectory of sleep disruptions in the acute period following a cardiac event.

## LIMITATIONS

Our study has several potential limitations. Using the PSQI as our primary metric for sleep health may have introduced recall bias. Future investigations should supplement the PSQI with objective sleep assessment, using wrist actigraphy for example. While we investigated changes in individual sleep health metrics and their associations with psychological distress, we did not capture novel sleep predictors of cardiometabolic health e.g., sleep regularity.^41^ Further, our examination of sleep patterns was confined to the initial month following a cardiac event. Recognizing that sleep dynamics may evolve, longitudinal studies with extended follow-up periods could delineate comprehensive sleep trajectories and elucidate their temporal interplay with psychological distress.

## CONCLUSIONS

We found significant sleep health deterioration among close family members one month after their loved one’s cardiac arrest. Cardiac arrest-induced psychological distress had a significant association with worsening in multiple aspects of sleep health. We found racial and ethnic disparities in these associations, with Hispanic families showing heightened susceptibility to sleep disruptions. Families prioritized interventions addressing information-based needs in the first month after cardiac arrest to reduce uncertainty and psychological distress. Our findings represent an initial important step in locating such factors in the conceptual model, considering age, sex, and race/ethnicity. Our research framework that integrates knowledge, methods, and measures from the fields of psychology and sleep research will catalyze advances in CVD prevention and more holistic management of critically ill patients, considering the secondary impact that their disease course may have on the psychological and cardiovascular health of close family members of critically ill patients. Further research is warranted to inform the development of comprehensive support strategies tailored to meet the multifaceted needs of families affected by cardiac arrest.

## Supplemental Material

Supplemental Methods

Supplementary Figure 1

Supplementary Tables 1-9

## Data Availability

The data that will support the findings of this study are available from the study investigators, but restrictions apply to the availability of these data, and so are not publicly available. Data are however available from the authors upon reasonable request and with permission of the study group.

